# Predictive Value of Blood Tests in Postoperative Delirium for Abdominal Surgery Patients

**DOI:** 10.64898/2026.03.04.26347627

**Authors:** Wesley Chorney, Michael Lisi

## Abstract

**Background:** Postoperative delirium is a common complication in surgical patients, and is associated with a multitude of negative outcomes, including mortality, dementia, and increased healthcare costs. Therefore, a better understanding of what factors contribute to postoperative delirium, especially those that can be easily obtained, is important.

**Methods:** We conducted a retrospective cohort study using patients from the Medical Information Mart for Intensive Care (MIMIC)-IV database. Adult patients undergoing procedures in abdominal surgery who did not have pre-existing delirium were included in the study. Overall, we included 8022 procedures across 7212 patients. For each admission, we extracted values obtained from common blood tests, the Charlson and Elixhauser comorbidity score, and patient demographic information. We used stepwise logistic regression to identify predictive factors of postoperative delirium in this cohort.

**Results:** The model isolated factors well known to be associated with postoperative delirium, such as age, comorbidity (as represented by the Elixhauser comorbidity score), and Parkinson’s disease. The model also selected variables that are less studied, such as minimum preoperative platelets and maximum preoperative sodium levels. We hypothesize that the former is associated with postoperative delirium as a surrogate marker for inflammation as an acute phase reactant, and the second due to it being a marker for cerebral edema and altered neurotransmission.

**Conclusion:** Preoperative blood tests contain valuable information that can be used alongside patient demographics and past medical history to better predict the risk of postoperative delirium.

## 1. Introduction

Delirium is a complex state of acute cerebral dysfunction characterized by acute onset, confusion, altered mental status, and fluctuating course [1]. In elderly patients, a recent meta analysis estimated an incidence of 24% across all types of surgeries, and 23% for noncardiac surgery [2]. Postoperative delirium (POD) is associated with a host of negative outcomes, including dementia and executive dysfunction [3], mortality [4], increased healthcare costs [5], and increased odds of non-home discharge [6]. Because delirium is a complex process involving an interaction between many precipitating and predisposing factors [7], it can be difficult to predict which patients are at high risk. However, it is estimated that a significant portion of cases are preventable [8], therefore, understanding those factors that contribute to POD is an important task, and being able to optimize at-risk patients prior to surgery could result in better outcomes and reduced healthcare costs.

There are a number of factors currently known to be independent predictors of postoperative delirium. A recent analysis showed that higher number of comorbidities, American Society of Anesthesiologists status III or IV, male sex, lower educational level, smoking, history of delirium, older age, history of institutionalization, being underweight, and longer duration of surgery were associated with the development of postoperative delirium in noncardiac surgery patients [9]. Other work has shown that preoperative use of opioids or benzodiazepines, preoperative infection, and hematocrit *<* 30% were also associated with the development of postoperative delirium [10]. Perioperative hypotension has also been noted as a risk factor for postoperative delirium in noncardiac surgery patients [11]. Preoperative stroke and high day one postoperative cortisol are also known to be risk factors [12]. Additional factors associated with POD include depression [13], dementia [14], alcohol use [15, 16], and Parkinson’s disease [17]. High preoperative C-reactive protein is also known to be associated with POD [18].

While these factors are associated with POD, only a subset of them are suitable for preoperative optimization. For instance, a high preoperative C-reactive protein may prompt additional investigation into infective or autoimmune processes in a patient, which may lead to proper treatment and optimization of this parameter prior to surgery; however, the presence of dementia or Parkinson’s disease are factors that cannot be optimized and are purely prognostic in nature. Therefore, additional studies are warranted on this topic. In particular, the present study investigates a large cohort of patients undergoing abdominal surgery, in conjunction with a number of readily available laboratory values, to ascertain which are prognostic of post-operative delirium. Clear relationships are shown, by means of logistic regression, between laboratory values and their prognostic value, which in turn sheds light on how these values may be optimized prior to surgery.

## 2. Materials and Methods

### 2.1. Data Source

We conducted a retrospective cohort study using the Medical Information Mart for Intensive Care IV (MIMIC-IV), version 3.1, a large, single-center, deidentified database containing detailed clinical data for patients admitted to the Beth Israel Deaconess Medical Center between 2008 and 2019 [19, 20], available from PhysioNet [21]. MIMIC-IV includes demographics, diagnoses, procedures, laboratory measurements, medications, and outcomes for both ward and intensive care unit (ICU) hospitalizations. Use of the database was approved under the data use agreement, and the study was exempt from institutional review board oversight due to the use of fully deidentified data.

Adult patients (defined as those 18 years of age or older) who underwent general surgical procedures during a hospital admission were identified. Surgical procedures to be included were defined via Internaional Classification of Diseases (ICD)-9 procedure codes and ICD-10 procedure codes, and were chosen to encompass common general surgery operations such as cholecystectomy, hernia repair, bowel resection, and related abdominal procedures. For patients with multiple surgical procedures during the same hospitalization, procedures were collapsed to the earliest qualifying surgical date, which was treated as the index operation. Patients were excluded if there was evidence of pre-existing delirium prior to the index surgery, defined by any delirium diagnosis code (either ICD-9 or ICD-10) recorded before the index operation.

The primary exposure was undergoing a qualifying general surgical procedure. Surgical urgency was classified as elective versus emergency using admission type and procedure context, with emergency surgery defined by non-elective admissions or procedures occurring in the setting of urgent or emergent hospital admission. Patient age was calculated at the time of surgery. Sex was obtained from the admissions and patients tables. Baseline comorbidity burden was also calculated using the Charlson [22] and Elixhauser [23] comorbidity indices (CCI and ECI, respectively), calculated at the admission level. Elixhauser comorbidities were derived from ICD-10 diagnosis codes where possible using the van Walraven weights [24] or from ICD-9 diagnosis codes using the Quan weights [25] where ICD-10 codes were not available. BMI was included based on height and weight obtained from the first-day derived anthropometric tables in MIMIC-IV.

Preoperative laboratory values were extracted from hospital laboratory records. For each patient, laboratory tests obtained within 7 days prior to the index surgery and up to the time of surgery were included. We included hematologic laboratory values (white blood cell count, hemoglobin, hematocrit, red blood cell count, platelet count), renal and electrolyte values (creatinine, blood urea nitrogen, sodium, potassium, chloride, bicarbonate), and hepatic values (total bilirubin). For each laboratory parameter, the minimum and maximum values within the preoperative window were calculated and included as separate covariates, as these have been shown to carry prognostic value with respect to mortality for other types of surgery [26, 27].

The primary outcome was the development of postoperative delirium within 30 days of the index surgical procedure. Delirium was identified using ICD-9 and ICD-10 codes consistent with acute confusional states and delirium syndromes recorded during the postoperative period. Patients with delirium documented prior to surgery were excluded.

### 2.2. Statistical Analysis

A stepwise multivariable logistic regression model was used for determining which factors were linked to postoperative delirium. A significance level of *α*_*i*_ = 0.03 was required for inclusion in the model, and a significance level of *α*_*r*_ = 0.05 was required for a variable to remain included in the model. Logistic regression was built using *statsmodels* version 0.14.4 [28] in Python 3.13.5 [29]. Groups in the study were defined by the development of post-operative delirium within 30 days of the surgery index date. Descriptive statistics were computed between the groups. Comparisons of continuous variables between groups were made with Welch’s two-sided *t*-test, with a significance level of *α* = 0.05. Categorical variables were compared with a chi-square test.

## 3. Results

Table 1 gives an overview of the dataset with respect to each variable. The *p*-values displayed are univariate analyses, and therefore, we not all of the variables with statistically significant *p*-values will be present in the logistic regression models due to co-linearity. Nevertheless, the strongest factors associated with postoperative delirium were both the Charlson comorbidity index and the Elixhauser score, as demonstrated in [30], age at surgery, and minimum and maximum preoperative total bilirubin.

**Table 1:** Variable distribution between the two groups, defined by post-operative delirium of surgery index date. For continuous variables, mean plus or minus standard deviation is displayed; whereas counts (with percentages in brackets) are displayed for categorical variables. Female sex is coded as 1 and male sex as 0.

| Variable | No Delirium | Delirium | p-value |
| --- | --- | --- | --- |
| N | 7934 | 92 |  |
| Age At Surgery | 60.2 ± 16.3 | 69.4 ± 15.3 | 0.000 |
| Charlson Comorbidity Index | 4.2 ± 3.2 | 5.3 ± 2.9 | 0.000 |
| Elxhauser Score | 6.5 ± 7.5 | 9.3 ± 6.5 | 0.000 |
| RBC Minimum | 3.6 ± 0.8 | 4.5 ± 0.9 | 0.005 |
| RBC Maximum | 4.0 ± 0.7 | 4.5 ± 0.9 | 0.048 |
| WBC Minimum | 8.1 ± 5.1 | 12.8 ± 4.1 | 0.004 |
| WBC Maximum | 11.1 ± 7.4 | 12.8 ± 4.1 | 0.202 |
| Hemoglobin Minimum | 10.6 ± 2.3 | 13.9 ± 3.2 | 0.006 |
| Hemoglobin Maximum | 11.6 ± 2.1 | 13.9 ± 3.2 | 0.041 |
| Hematocrit Minimum | 32.5 ± 6.7 | 42.0 ± 8.3 | 0.004 |
| Hematocrit Maximum | 35.6 ± 5.9 | 42.0 ± 8.3 | 0.029 |
| Platelets Minimum | 209.5 ± 105.1 | 314.7 ± 130.1 | 0.023 |
| Platelets Maximum | 249.3 ± 119.7 | 314.7 ± 130.1 | 0.126 |
| Sodium Minimum | 136.6 ± 4.4 | 138.1 ± 4.8 | 0.337 |
| Sodium Maximum | 139.9 ± 4.1 | 138.1 ± 4.8 | 0.273 |
| Potassium Minimum | 3.8 ± 0.5 | 4.3 ± 0.5 | 0.024 |
| Potassium Maximum | 4.4 ± 0.7 | 4.5 ± 0.8 | 0.798 |
| Creatinine Minimum | 1.0 ± 1.0 | 1.3 ± 0.7 | 0.149 |
| Creatinine Maximum | 1.3 ± 1.4 | 1.3 ± 0.7 | 0.753 |
| BUN Minimum | 15.7 ± 13.1 | 29.0 ± 22.8 | 0.099 |
| BUN Maximum | 21.8 ± 18.9 | 29.0 ± 22.8 | 0.346 |
| Bilirubin Total Minimum | 2.5 ± 4.6 | 0.9 ± 0.6 | 0.000 |
| Bilirubin Total Maximum | 3.5 ± 6.2 | 0.9 ± 0.6 | 0.000 |
| Albumin Minimum | 3.3 ± 0.7 | 3.7 ± 0.6 | 0.088 |
| Albumin Maximum | 3.5 ± 0.7 | 3.7 ± 0.6 | 0.328 |
| Sex | 3832 (48.3%) | 49 (53.3%) | 0.34748 |
| Alcohol Use | 418 (5.3%) | 8 (8.7%) | 0.15456 |
| Depression | 1159 (14.6%) | 19 (20.7%) | 0.10430 |
| Parkinson's | 41 (0.5%) | 5 (5.4%) | 0.00017 |
| Dementia | 79 (1.0%) | 3 (3.3%) | 0.06767 |
| Elective Surgery | 225 (2.8%) | 8 (8.7%) | 0.00511 |

The logistic regression model is fit using ten-fold stratified cross validation, and in model training, the no delirium class is downsampled to be thrice as numerous as the delirium class. Predictors are tracked over each fold and the frequency with which they are selected over the ten folds is displayed in Table 2, which also displays odds ratios (ORs) for each coefficient.

**Table 2:** Coefficients reported as odds ratios, as well as their selection frequency across folds. 95% confidence intervals for each odds ratio are also reported.

| Predictor | Selection Frequency (%) | Folds Included | Mean OR | OR 2.5% | OR 97.5% |
| --- | --- | --- | --- | --- | --- |
| Age | 100 | 10 | 1.046193 | 1.034668 | 1.062051 |
| Elixhauser Score | 100 | 10 | 1.099381 | 1.067151 | 1.135486 |
| Maximum Preoperative Sodium | 90 | 9 | 0.937165 | 0.894856 | 0.976290 |
| Minimum Preoperative Platelets | 70 | 7 | 1.008562 | 1.006221 | 1.011700 |
| Elective Surgery | 60 | 6 | 6.023315 | 4.423827 | 10.677724 |
| Parkinson's Disease | 50 | 5 | 24.973628 | 8.419039 | 142.340699 |
| Minimum Preoperative Hematocrit | 40 | 4 | 1.567265 | 1.209215 | 1.925828 |
| Minimum Preoperative Hemoglobin | 30 | 3 | 6.047791 | 4.968936 | 7.382581 |
| Charlson Comorbidity Index | 30 | 3 | 0.846931 | 0.832353 | 0.858129 |
| Maximum Preoperative Hematocrit | 30 | 3 | 0.559571 | 0.505133 | 0.614753 |
| Minimum Preoperative Potassium | 20 | 2 | 5.527832 | 5.156696 | 5.925680 |
| Maximum Preoperative Red Blood Cells | 20 | 2 | 0.007719 | 0.007170 | 0.008311 |
| Alcohol Use | 10 | 1 | 3.583869 | 3.583869 | 3.583869 |
| Minimum Preoperative Albumin | 10 | 1 | 5.716050 | 5.716050 | 5.716050 |

We note that age and Elixhauser score were selected for inclusion in every fold, maximum preoperative sodium was selected in nine of ten folds, and both minimum preoperative platelets and elective surgery were selected for inclusion in over half of all folds. Generally, increased frequency of selection signifies a stronger association to the outcome (postoperative delirium), over and above other variables which may be associated, but are collinear with, a selected variable. For instance, there is significant collinearity between ECI and CCI, and indeed, we see that CCI was selected for inclusion in only three folds. Odds ratios vary significantly between coefficients, with consistently selected variables typically displaying modest ORs with narrow confidence intervals (CIs); whereas variables such as elective surgery and Parkinson’s disease have high ORs with wide CIs.

Table 3 and Figures 1 and 2 describe the model performance. The model is evaluated with respect to area under the receiver-operator characteristic curve (ROC-AUC), area under the precision recall curve (PR-AUC), accuracy, precision, sensitivity, specificity, and F1 score (which is the harmonic mean of precision and sensitivity). All of these metrics range from zero to one. Generally, the logistic regression model is accurate, specific, and has a decent ROC-AUC score, but some cases of postoperative delirium go undetected (as evidenced by the low precision, sensitivity, PR-AUC, and F1 score). Figure 1 displays the ROC with a bootstrapped 95% confidence interval, as well as the performance of a random chance classifier (the dashed line), which would have an ROC-AUC of 0.5. We note the model performs significantly better than this. Figure 2 displays the PR curve, again with a bootstrapped 95% confidence interval. The regression models is compared to a classifier that outputs the raw probability of post-operative delirium in the dataset, and performs significantly better than this.

**Table 3:** Model metrics across the ten folds. For each metric, the mean value, as well as the lower and upper bounds of a 95% CI are displayed.

| **Metric** | **Mean** | **CI Lower (2.5%)** | **CI Upper (97.5%)** |
| --- | --- | --- | --- |
| ROC-AUC | 0.779 | 0.739 | 0.820 |
| PR-AUC | 0.038 | 0.026 | 0.059 |
| Accuracy | 0.931 | 0.925 | 0.936 |
| Precision | 0.042 | 0.025 | 0.061 |
| Sensitivity | 0.228 | 0.144 | 0.316 |
| Specificity | 0.939 | 0.934 | 0.944 |
| F1 Score | 0.070 | 0.043 | 0.102 |

**Figure 1.**
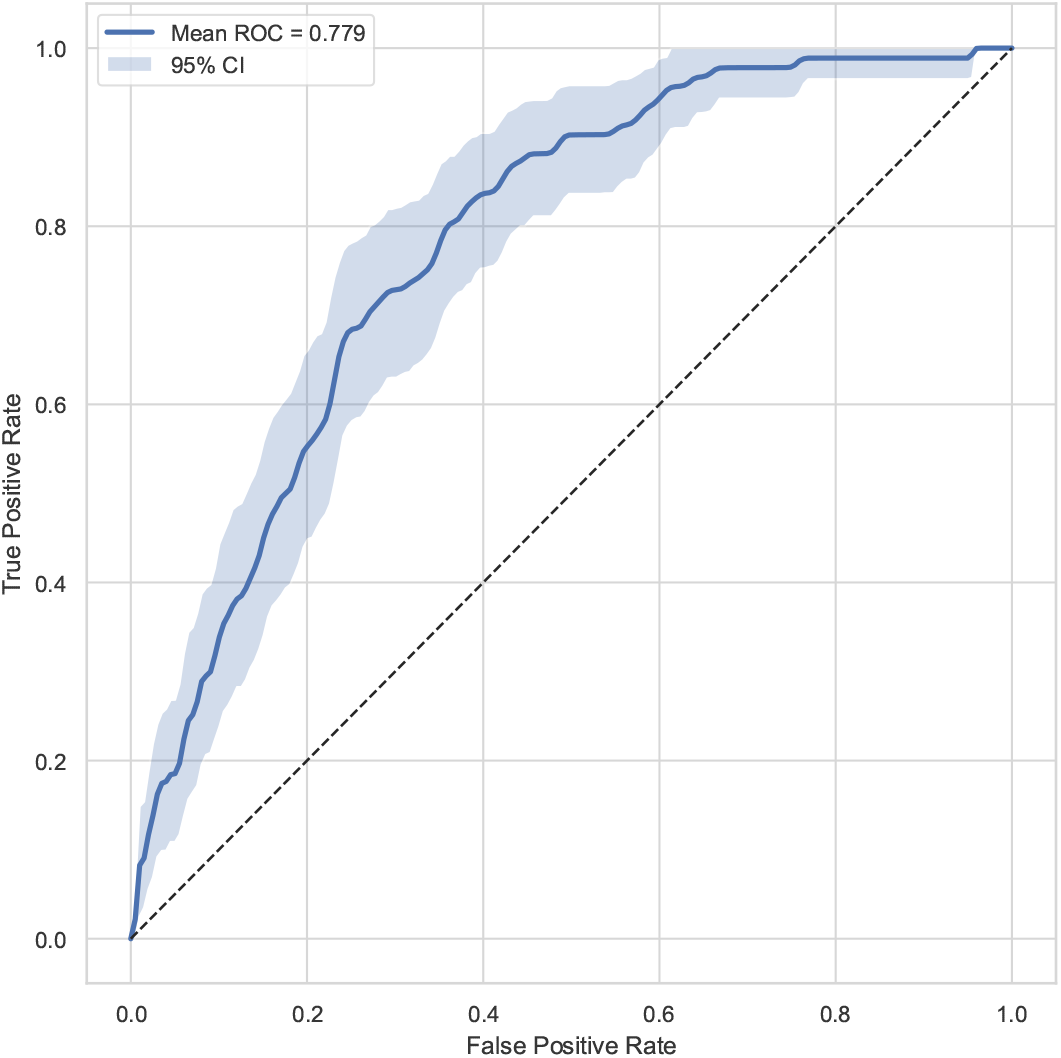
AUC-ROC curve for the model, with bootstrapped 95% confidence interval.

**Figure 2.**
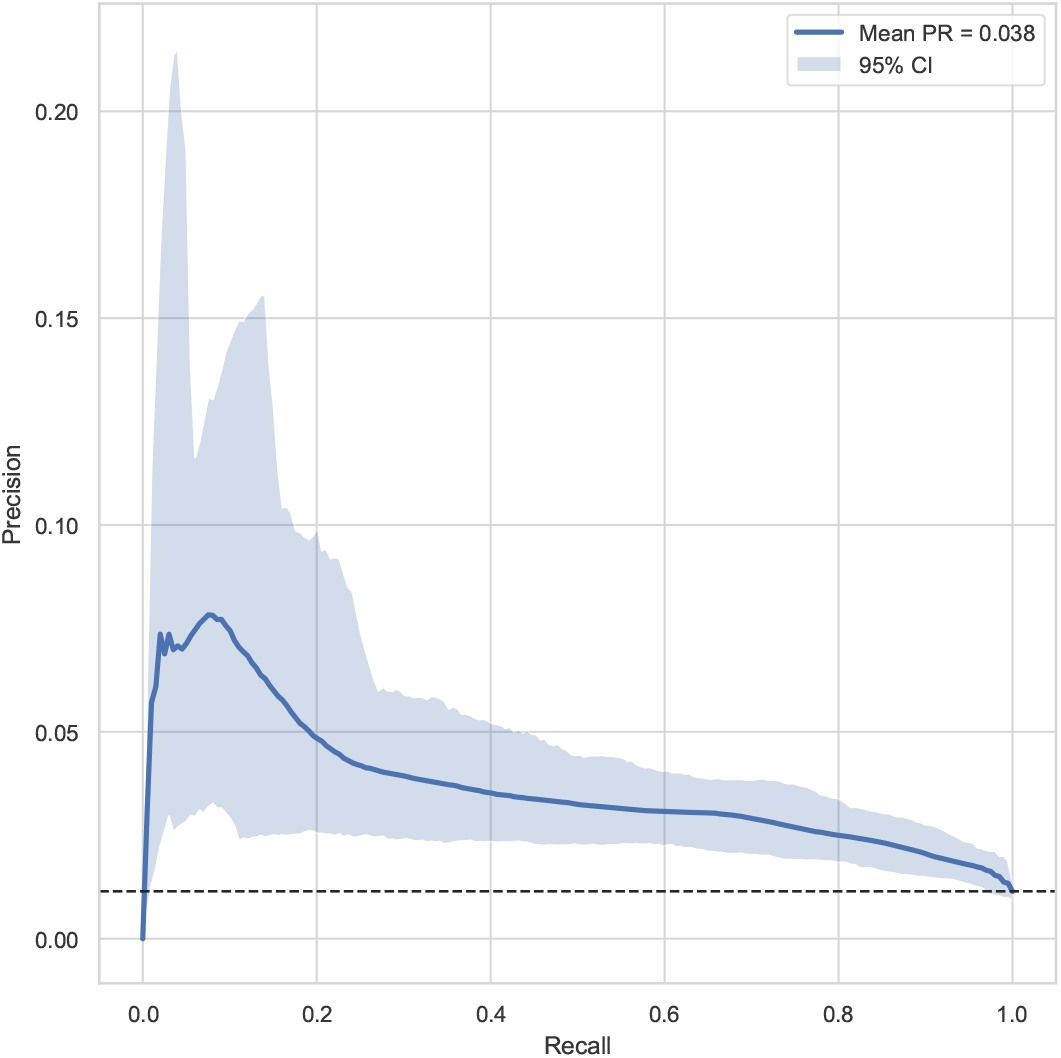
AUC-PR curve for the model, with bootstrapped 95% confidence interval. Note the *y*-axis is scaled for visibility.

## 4. Discussion

On the basis of a univariable analysis displayed in Table 1, many variables are statistically significantly associated with POD. However, there is likely to be collinearity in the raw data. For instance, the CCI and ECI are collinear, and some laboratory values, such as red blood cells, hematocrit, and hemoglobin, are likely to be collinear, at least in a subset of cases. The use of a multistep logistic regression model allows for controlling of collinearity and confounding by other variables at each step in the selection process, and for removal of variables that become insufficiently associated with the outcome upon inclusion of other variables. Therefore, we see only a subset of these variables frequently included in the regression model across all ten folds.

Variables that are known to correlate with POD are frequently selected for inclusion into the model. For instance, both age and ECI are known to be associated with postoperative delirium [31, 30]. Parkinson’s disease is also known to be associated [17]. This, in conjunction with strong model performance as evidenced by the AUC-ROC score, accuracy, and specificity, is reassuring in that the model aligns well with existing studies. It also indicates that the dataset is most likely consistent with other clinical datasets that have been used in investigating similar questions, and while further work would be required to ensure external validity of the present model, it is encouraging that factors known to be associated with postoperative delirium are also highlighted by the present study.

However, we also observed that maximum preoperative sodium was negatively associated with postoperative delirium. Those patients lacking high preoperative sodium had a consistently low (or lower than average) sodium level. While the difference between groups in Table 1 is not statistically significant, it is possible that there is a subset of patients with significantly lower levels for whom this is a strong association. While postoperative hyponatremia is known to be associated with delirium [32], and preoperative hyponatremia is known to be associated with postoperative delirium in coronary artery bypass grafting in elderly patients [33], it is not well-studied as a causative factor for postoperative delirium. Nevertheless, given that hyponatremia is associated with delirium in general [34], and that low sodium can contribute to cerebral edema, raised intracranial pressure, and disruption to astrocyte volume regulation and neurotransmission [35], it is likely that low maximum preoperative sodium is a marker for sustained hyponatremia, or sustained subclinical hyponatremia, which could precipitate these effects. Ultimately, this emerges as a clear variable which should be optimized prior to abdominal surgery.

Similar to the above, higher levels of minimum preoperative platelets were associated with the development of POD. Inversely to maximum preoperative sodium, a higher value of minimum preoperative platelets is achieved when platelets are consistently elevated. As an acute phase reactant, this may be a marker of inflammation, which is well-known to correlate with the development of delirium [35, 36]. While the bioenergetic health index of platelets is known to be associated with postoperative delirium [37], and thrombocytosis itself is known to be associated with a number of postoperative conditions, but not delirium [38], the current study highlights the predictive value of elevated minimum preoperative platelets wit respect to POD. This is another variable which should be optimized prior to abdominal surgery.

We note that the incidence of postoperative delirium in our dataset was approximately 1%, which differs from estimates in the literature — for instance, a large cohort study of elderly patients reported an incidence of approximately 10% [39]. It is worth noting that the inclusion criteria in the aforementioned study versus the previous study differ significantly. The present study allows for a broad age range of patients undergoing both elective and non-elective abdominal surgery, whereas the aforementioned study focused on elderly adults undergoing elective abdominal surgical procedures. More generally, the incidence of postoperative delirium in noncardiac surgery in an adult population is estimated to be 17.7% [9]; however, this study considers any procedures that are not cardiac and does not focus specifically on abdominal surgery. Nevertheless, the incidence in our dataset is lower than expected, and further investigation could seek to what extent the factors that we isolated remain predictive factors in other datasets. Another limitation is that logistic regression models cannot adequately capture interaction between variables. Deep learning methods can model higher-order interactions between features, and would allow for the detection of more complex pattern between variables; however, this comes at the cost of explainability. While logistic regression cannot model interactions between variables, it can directly quantify the effect of a given variable amongst a set of variables on an outcome.

## 5. Conclusion

Postoperative delirium is an important complication of abdominal surgery. It is associated with increased morbidity, mortality, and healthcare burden. While many studies have investigated POD with respect to certain procedures and a geriatric population, we sought to investigate those factors associated with POD in a broader population undergoing common procedures in abdominal surgery. We used a dataset containing 8,022 procedures from 7,212 patients in the MIMIC-IV database, and applied stepwise logistic regression with stratified cross fold validation to determine predictors of postoperative delirium in this cohort. Our model was well aligned with existing literature, highlighting factors such as age, comorbidity (as captured by the ECI), and Parkinson’s disease. However, we also identified factors not commonly described in the literature, including maximum preoperative sodium and minimum preoperative platelets. We described how these could be related to postoperative delirium (relating to cerebral edema and neurotransmission, and inflammatory states, respectively).

There are two immediately salient areas for future studies to focus on. First, it may be worth expanding our approach to other clinical datasets and determining to what extent these results hold. In the context of maximum preoperative sodium and minimum preoperative platelets, there is a clear mechanism through which we expect post-operative delirium is caused, but it would be helpful to observe whether this association is observed in other datasets. Second, other modelling approaches could be used. While deep learning methods offer suffer from a lack of explainability, it is possible that such methods would better detect potential cases of POD, and methods from explainable artificial intelligence could be used to derive further insights.

## Data Availability

The data analyzed in this study were obtained from the MIMIC-IV database (version 3.1). Access to MIMIC-IV requires completion of human subjects research training and data use agreement through PhysioNet. Due to data use restrictions, the raw clinical data cannot be publicly shared by the authors, but qualified researchers may request access directly from PhysioNet (https://physionet.org/content/mimiciv/3.1/) under the same terms.

